# Identifying secondary findings in PET/CT reports in oncological cases: A quantifying study using automated Natural Language Processing

**DOI:** 10.1101/2022.12.02.22283043

**Authors:** Julia Sekler, Benedikt Kämpgen, Christian Philipp Reinert, Andreas Daul, Brigitte Gückel, Helmut Dittmann, Christina Pfannenberg, Sergios Gatidis

## Abstract

**Background:** Because of their accuracy, positron emission tomography/computed tomography (PET/CT) examinations are ideally suited for the identification of secondary findings but there are only few quantitative studies on the frequency and number of those.

Most radiology reports are freehand written and thus secondary findings are not presented as structured evaluable information and the effort to manually extract them reliably is a challenge. Thus we report on the use of natural language processing (NLP) to identify secondary findings from PET/CT conclusions.

**Methods:** 4,680 anonymized German PET/CT radiology conclusions of five major primary tumor entities were included in this study. Using a commercially available NLP tool, secondary findings were annotated in an automated approach. The performance of the algorithm in classifying primary diagnoses was evaluated by statistical comparison to the ground truth as recorded in the patient registry. Accuracy of automated classification of secondary findings within the written conclusions was assessed in comparison to a subset of manually evaluated conclusions.

**Results:** The NLP method was evaluated twice. First, to detect the previously known principal diagnosis, with an F1 score between 0.65 and 0.95 among 5 different principal diagnoses.

Second, affirmed and speculated secondary diagnoses were annotated, and the error rate of false positives and false negatives was evaluated. Overall, rates of false-positive findings (1.0%-5.8%) and misclassification (0%-1.1%) were low compared with the overall rate of annotated diagnoses. Error rates for false-negative annotations ranged from 6.1% to 24%. More often, several secondary findings were not fully captured in a conclusion. This error rate ranged from 6.8% to 45.5%.

**Conclusions:** NLP technology can be used to analyze unstructured medical data efficiently and quickly from radiological conclusions, despite the complexity of human language. In the given use case, secondary findings were reliably found in in PET/CT conclusions from different main diagnoses.

## Background

In order to evaluate clinically relevant questions both retrospectively and prospectively within studies as well as for therapy optimization, it is often necessary to evaluate radiological reports since these are important sources of clinical diagnostic information. However, manual evaluation is only possible with a significant effort if a large number of reports and findings are involved (1). To extract important information from freehand texts, artificial intelligence applications can be helpful. However standardized artificial intelligence (AI) applications are difficult to establish, since radiological texts are usually freely written and language use and vocabulary are heterogeneous. Therefore, particular AI solutions are needed, such as natural language processing (NLP). These can evaluate certain questions quickly, effectively and error-controlled and can be adapted to the respective problem.

NLP describes a subfield of AI. It is used in numerous medical applications where text data has to be analyzed and human writing or speech has to be understood and interpreted. For example, this includes medical chatbots, in retrospective selection of data from unstructured records (2), in research queries (3) in billing and coding (4), and in studies to analyze drug safety (5, 6).

Regarding the use of NLP for assessment of radiological reports, different clinical question have been addressed in previous studies such as the detection of suspicious findings in mammography (7), identification of site-specific bone fractures (8), tumor stage NLP (9) and other specified diagnoses (10, 11) (12, 13). In this context, NLP is increasingly being used for the extraction of relevant information from radiology reports in clinical studies (14-16).

PET/CT is mainly used for detection of tumor lesions and staging of tumor spread in oncological patients. Major tumor entities examined by PET/CT include Melanoma, Prostate Cancer, Lung Cancer, Lymphoma and Neuroendocrine Tumors (17-21). However, not only the status of the known or suspected disease is crucial for therapy and patient management but also clinically relevant secondary findings such as inflammation, vascular complications and unknown secondary tumors. Incidental findings are quite common (22) and can be important for definition of scan protocols, reporting strategies and further management, especially in oncology.

The purpose of this study was thus to automatically extract information about the occurrence of secondary findings by automated analysis of freehand written radiological conclusions using NLP.

## Methods

This study was based on a PET/CT registry (04/2013 – 12/2018) (23) including 7715 scans in total. The study was reviewed and approved by the local institutional review board (Ethics committee of the University of Tuebingen, reference number 064/2013B01). Informed consent regarding the use of data for research was obtained from all patients.

### PET/CT protocols

All PET/CT examinations were performed on a state-of-the art clinical scanner (Biograph mCT, Siemens Healthineers, Knoxville, TN). using a standardized examination protocol. Different PET tracers were applied: [68Ga]-HA-DOTATATE in case of neuroendocrine tumors, [68Ga]-PSMA in case of prostate cancer, [11C]-Choline in case of prostate cancer, and [18F]-FDG in all other oncological indications. All CTs were acquired in full-dose technique with contrast agent where appropriate.

### Structure of reports

Free text PET/CT reports were written in German in a clinical routine setting using a standardized structure described as follows:

#### 1. Clinical Information

After providing an appropriate indication for the study by the referring physicians, the primary clinical questions to be answered by the PET/CT examination are documented in the reports.

#### 2. Technique

This section describes how the study was generated including information on the radiopharmaceutical used, the administered activity and the CT technique. Also, the axial coverage of the scan was documented (e.g., “skull base to mid-thigh”). In certain cases, PET/CT protocols may have included additional acquisitions such as delayed imaging.

#### 3. Previous Studies

All reports included information on prior studies which are used for comparison or correlation. If no previous imaging studies are available, this was also stated.

#### 4. Findings

Findings were organized by anatomic region describing both PET and CT findings relevant to the clinical question within each anatomic subsection. This part also included a description of incidental PET and CT findings unrelated to the primary cancer being studied. The intensity of radiotracer uptake was reported using both qualitative (e.g. moderate or intense) terminology as well as semiquantitative measures such as the SUV.

#### 5. Conclusion

All reports concluded with a summarizing evaluation of the findings answering the specific clinical questions raised by the referring physician and providing a diagnosis or a brief list of differential diagnoses. In addition, potentially clinically relevant secondary findings were summarized in this section.

### NLP

The annotation of diagnoses in the report sections were automatically generated using a proprietary NLP tool, Empolis Knowledge Express by Empolis Information Management GmbH (Kaiserslautern, Germany; https://knowledge.express/). The Empolis NLP system (24, 25) implements a common NLP pipeline consisting of cleansing (e.g., replacement of abbreviations), contextualization (e.g. into segments “clinical information”, “findings”, and “conclusion”), concept recognition using common terminologies such as the Radiological Lexicon (RadLex) and the International Classification of Diseases (ICD), and negation detection (e.g., “affirmed”, “negated”, and “speculated”). The NLP system uses a neural language model and word embeddings trained with fastText (26) on a medical corpus of more than 100.000 German radiological reports and other medical literature (457 MB of text data). The language model computes for every word a 128-dimensional vector. For concept recognition, a full text index and morpho-syntactic operations such as tokenization, lemmatization, part of speech tagging, decompounding, noun phrase extraction and sentence detection were used. The index was populated with synonyms for all entities (both from terminologies and by manual extensions). For negation detection, typically, a rule-based approach is used (27); however, the heterogeneity in which pathological findings are affirmed, negated or speculated require a more elaborate learning approach. Therefore, the NLP system uses a bidirectional recurrent neural network based on two stacked Gated Recurrent Unit (GRU) layers (28) trained and validated on more than 2.000 manually labelled reports with negation information using the NLP library spaCy (29). Every input was a 50-word window, the output returned a negation status for each word. The validation dataset showed 0.93 accuracy. For the analysis by the Empolis NLP system, no pre-processing of the annotated radiological reports was necessary.

Findings identified by the NLP system were classified in two categories: Unconfirmed secondary findings, such as those given as differential diagnoses or as suspicions, were annotated as *speculated*, whereas confirmed diagnoses are annotated as *affirmed*.

For automated detection of the primary patient diagnosis, the *Clinical Information* field of the radiology report was used, for automated detection of secondary findings identified in the PET/CT examination, the *Evaluation* field was used as input to the NLP system.

### The Radiological Lexicon (RadLex)

In order to interpret radiological findings by NPL in a standardized way, a uniform representation of the radiological terms is required. The Radiological Lexicon (RadLex) was developed to standardize radiological terms (30). RadLex consists of a uniform vocabulary of radiological terminology that is organized hierarchically so that relationships between terms are maintained (31). In RadLex terminology there are very detailed terms for anatomy, pathology and radiological diagnoses. Some of these concepts, such as the diagnosis “neuroendocrine tumors”, are therefore much easier to map with the RadLex system compared to other coding systems, such as the ICD system.

### Annotation of radiological evaluations of PET/CT scans Selection of scans

A total of 4680 scans in patients with the 5 most frequent tumor entities from the registry was annotated in this study (melanoma, non-hodgkin-lymphoma (NHL), lung cancer (lung-CA), prostate cancer (prostate-CA) and neuroendocrine tumors (NET)). Only scans from patients investigated for staging in either histologically affirmed or speculated malignancy of the above-mentioned entities were allowed. Reports were anonymized to remove patient identifiers. All characteristics of chosen scans are listed in Table.

### Annotation of radiological conclusions

#### Annotation of clinical information

In order to estimate the performance of the NLP system in a setting with available ground truth, the primary diagnosis was annotated first. The system was supposed to find out the main or tentative diagnosis which is, in most cases, noted in the clinical information.

Since the principal diagnoses may be indicated with different synonyms or paraphrases within the clinical information, synonyms or paraphrases were introduced into the NLP system. Subsequently, the F1-score, positive predictive value and sensitivity were calculated.

#### Annotation of secondary findings

Only the conclusion and not the entire report was used for annotation of the main and secondary diagnoses.

All radiological evaluations were uploaded onto a healthcare-analytics database provided by Empolis Information Management GmbH. In this database all secondary findings, that were automatically annotated were presented in a structure analogous to RadLex (31) in which supersets were in turn subdivided into further specific subgroups. This categorization provides a hierarchical representation of diagnoses with more general supersets such as “infectious or inflammatory disease” as well as more specific subgroups such as “sinusitis”. Most secondary findings were categorized within these specific subgroups; remaining (rare) findings among the supersets were subsumed into the more general categories, such as “infectious or inflammatory disease” or “mechanical disorder” and will be referred to as “others” in the following. All affirmed or speculated secondary tumors, are subsumed as a separate category of supersets. These are not further divided subgroups.

A list of all annotated diagnoses and their division into supersets and subgroups with the corresponding RadLex codes can be found in the supplementary material (S1 Fig).

### Assessment of algorithm performance for classification of primary diagnosis

To assess algorithm performance for classification of the primary diagnosis, algorithm output derived from the clinical information field was compared to the actual clinical diagnosis of each patient. Accuracy, positive predictive value and sensitivity were computed.

### Assessment of algorithm performance for classification of secondary findings

For automated classification of secondary findings, algorithm output was compared to the content of the conclusion section of each radiological conclusion. To this end all findings generated by the algorithm were re-evaluated by two experts in medical imaging identifying correct and false positive findings. For the evaluation of false-positive findings, the number of false-positive findings was counted by manual verification by two experts in medical imaging. False positive findings were divided in two categories: Non-annotated finding or wrong level of uncertainty (speculated vs. affirmed).

All secondary findings in total were summarized and the percentage of false positives was calculated as a result. The number of false positives in which affirmed and speculated are interchanged was also analyzed.

In order to estimate the frequency of false negative findings, a random sample of 500 radiological conclusions (100 per cancer entity) were manually evaluated by two experts in medical imaging identifying secondary findings that were not captured by the NLP system. Subsequently, all manually recorded secondary findings were matched with those found by the NLP system.

### Statistical analysis

To evaluate the performance of the NLP system in detecting the principal diagnosis from the clinical information, we calculated the overall correlation between the proposed NLP algorithm and the gold standard. Three metrics, being sensitivity, specificity, and F1-score, were used for this purpose.

For the evaluation of the NLP system for annotation of secondary findings, false-positive and false-negative cases were counted and correlated to the total number of annotations.

## Results

### Quality of automated annotation

#### Classification of main diagnoses

The NLP system’s performance was first tested regarding the classification of the primary diagnosis. The system achieved an F1-score of 0.95 for the diagnosis of melanoma, 0.65 for the diagnosis of lung-CA, 0.90 for the diagnosis of prostate-CA, and 0.90 for the principal diagnosis of NHL showing the efficacy of the NLP system for identifying primary diagnoses from clinical information. The lowest F1-score with 0.65 was achieved for lung-CA. We achieved a perfect positive predictive value in melanoma, NHL and prostate-CA demonstrating that the NLP algorithm has high precision in identifying primary diagnoses from clinical information. The best sensitivity was in melanoma with 0.91 whereas we got the lowest sensitivity with 0.49 in cases with lung-CA meaning that the system was able to identify between 49% and 91% of the cases. All primary diagnoses and the number of histologically affirmed and speculated cases with the respective F1-scores of the clinical information annotation are listed in Table 2.

**Table 1.** List of all scans, divided into the five tumor types with detailed characteristics.

|  |  | Melanoma | Prostate-CA | Lung-CA | NHL | NET |
| --- | --- | --- | --- | --- | --- | --- |
| **Total** |  | 1178 | 1255 | 928 | 533 | 786 |
| **(H. aff./ H. spec.)** |  | (1178/0) | (1255/0) | (901/27) | (533/0) | (719/67) |
| **Gender** | **Male** | 687 | 1255 | 627 | 314 | 417 |
|  | **Female** | 491 | 0 | 301 | 219 | 369 |
| **Age** |  | 63 (17 – 95) | 70 (44 – 88) | 66 (28 – 89) | 58 (6 – 87) | 62 (14 – 91) |

H. aff. = histologically affirmed H. spec. = histologically speculated
Age is presented as mean of years and range in parenthesis.

**Table 2.** Results of the annotation of the primary diagnosis in the clinical information of all scans.

|  | RadLex Code | *n*<br>H. aff. | *n*<br>H. spec. | Pos. pred.<br>value | Sensitivity | F1-Score |
| --- | --- | --- | --- | --- | --- | --- |
| **Lung-CA** | RIDE2220 Mass or nodule | 901 | 27 | 0.98 | 0.49 | 0.65 |
| **Melanoma** | RID34617 Melanoma | 1178 | 0 | 1 | 0.91 | 0.95 |
| **NET** | RID4483 Neuroendocrine neoplasm | 719 | 67 | 0.96 | 0.52 | 0.67 |
| **NHL** | RID3840 Lymphoma or<br>RID3843 non-Hodgkin lymphoma | 533 | 0 | 1 | 0,82 | 0,90 |
| **Prostate-CA** | RID45689 Prostate cancer | 1255 | 0 | 1 | 0.82 | 0.90 |

Pos. pred. value = Positive predictive value H. aff. = histologically affirmed H. spec. = histologically speculated
Pos. pred. value, sensitivity and F1-Score of the annotation of the h. aff. and spec. main diagnosis in the clinical information of all scans.

#### Classification and distribution of secondary findings

First, all secondary findings were combined into supersets to determine their distribution. Although distributions were quite similar within the main diagnoses, there were obvious differences (Figure 1). In general, the rate of “mechanical disorders” was highest in all cohorts but patients with lung CA had a very high rate of “mechanical disorders, comparatively.” This superset included subgroups such as atelectasis, thrombosis, and pleural effusion “Infectious or inflammatory disorders” such as pneumonitis, diverticulitis, and sinusitis occurred most frequently in patients with melanoma.

**Figure 1.**
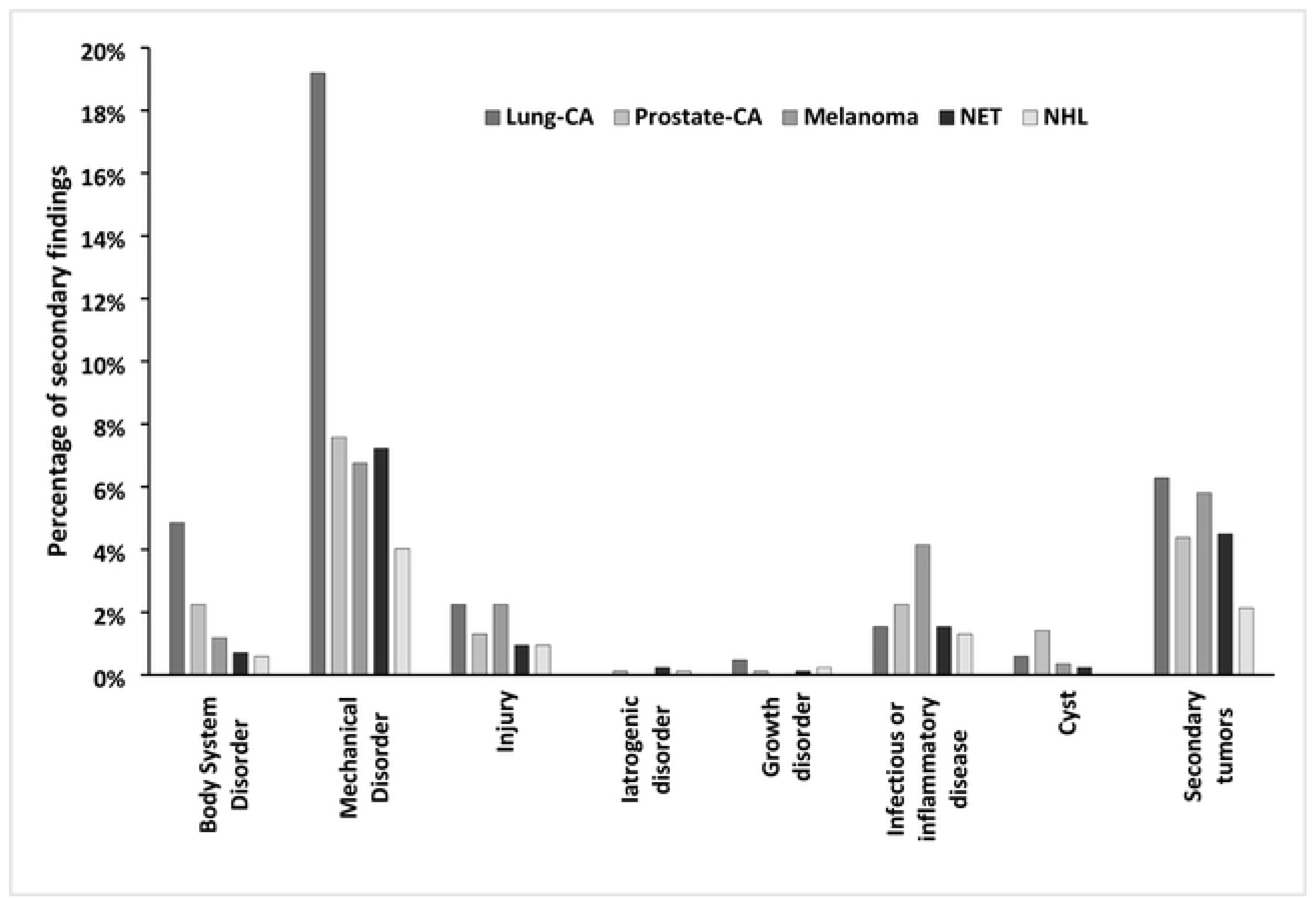
Distribution of affirmed supersets of secondary findings of all cohorts as identified by the NLP-System.

Second, supersets were divided into more specific subgroups (SG), secondary tumors (ST) and “others”, and their respective numbers were determined. “Others” included all secondary findings in supersets that were not specifically divided into further subgroups based on the RadLex hierarchy (Table 3).

**Table 3.** List of all superset categories with their included diagnoses.

| Superset category | Included diagnoses (=subgroups) |
| --- | --- |
| Body-system-specific disorder | Dissection, aneurysm, pseudoaneurysm, pneumonia, pneumothorax |
| Degenerative disorder | Degeneration*<sup>1</sup>, necrosis, deposition, ossification, resorption |
| Mechanical disorder | Atelectasis, thrombosis, embolism, pleural effusion, pericardial effusion, ascites, lymphocele, hernia, obstructive uropathy, hydronephrosis, hemorrhage, gallstone, urolithiasis, thrombus |
| Injury | Fracture |
| Iatrogenic disorder | Postoperative complication |
| Growth disorder | Cirrhosis |
| Infectious or inflammatory disease | Cholangitis, cholecystitis, diverticulitis, colitis, sarcoidosis, Crohn disease, ulcerative colitis, pancreatitis, pneumonitis, abscess, sinusitis |
| Cyst | Cyst*<sup>2</sup>, epidermoid, mucocoele |

List according to the RadLex hierarchy \*<sup>1</sup>http://radlex.org/RID/RID5043, \*<sup>2</sup>http://radlex.org/RID/RID3890)

The most affirmed specific subgroups in total were found in the cohort with the principal diagnosis of lung-CA (244 SG and 53 ST). This was followed in descending order by the cohorts with melanoma (124 SG and 49 ST), prostate-CA (127 SG and 37 ST), NET (93 SG and 38 ST), and the lowest number of subgroups was observed in patients with the principal diagnosis of NHL (61 SG and 18 ST). A differentiated analysis of the individual subgroups showed that this distribution occurred for almost all main diagnoses. Only “infectious or inflammatory diseases” occurred more frequently in melanoma patients than in all other. In particular, the secondary diagnosis “sinusitis” was found very often in this cohort. The greatest amount of “others” was identified in the cohort with lung CA (351). The lowest number was observed in the NET cohort (91). All results of the analysis are presented in Table 4 and Figure 2. The detailed distribution of all secondary findings can be found in the supplementary material S1 Fig.

**Table 4.** Distribution of affirmed and speculated subgroups and “others” in radiological conclusions.

|  |  | Lung-CA | Prostate-CA | Melanoma | NET | NHL |
| --- | --- | --- | --- | --- | --- | --- |
| **Total SG/SM (%)** | affirmed | 244/53<br>(9%/2%) | 127/37<br>(5%/1%) | 124/49<br>(5%/2%) | 93/38<br>(4%/1%) | 61/18<br>(2%/1%) |
|  | speculated | 53/71<br>(2%/3%) | 37/54<br>(1%/2%) | 49/51<br>(2%/2%) | 38/27<br>(1%/1%) | 18/22<br>(1%/1%) |
| **Total “others” (%)** | affirmed | 351 (13%) | 227 (9%) | 330 (13%) | 91 (3%) | 136 (5%) |
|  | speculated | 143 (5%) | 50 (2%) | 161 (6%) | 29 (1%) | 68 (3%) |

**SG = subgroups SM = secondary malignancies**
**Secondary malignancies (SM) are a special subgroup. Percentage in relation of the total number of secondary findings is shown in parentheses.**

**Table 5.** Calculation of false-positives.

| FP | Lung-CA | Prostate-CA | Melanoma | NET | NHL |
| --- | --- | --- | --- | --- | --- |
| Total SF<br>(aff./spec.) | 855<br>(648/207) | 531<br>(391/140) | 695<br>(503/192) | 295<br>(222/73) | 309<br>(215/94) |
| subgroups/"others" *n* | 425/430 | 267/264 | 274/421 | 176/119 | 131/178 |
| FP incorrect assignment<br>aff./spec. *n* (%) | 3 (0.4%) | 6 (1.1%) | 1 (0.1%) | 1 (0.3%) | 0 (0.0%) |
| FP incorrect SF *n* (%) | 14 (1.6%) | 27 (5.1%) | 8 (1.2%) | 17 (5.8%) | 3 (1.0%) |

**FP = false-positives SF = secondary findings aff. = affirmed spec. = speculated SF = secondary findings** **Results are presented as number and percentage (%) of total in parenthesis. Distinction was made** **between incorrect assignment of affirmed and speculated and incorrect SF. The percentage of total** **was calculated.**

**Table 6.** Calculation of false-negatives from a sample of 100 random radiological conclusions per principal diagnosis.

| FN | Lung-CA | Prostate-CA | Melanoma | NET | NHL |
| --- | --- | --- | --- | --- | --- |
| Conclusions with SF | 45 | 25 | 19 | 33 | 44 |
| Non-annotated SF *n* (%) | 5 (11.1%) | 6 (24.0%) | 2 (10.5%) | 2 (6.1%) | 8 (18.2%) |
| Deficit multiple SF *n* (%) | 9 (20.0%) | 7 (28.0%) | 5 (26.3%) | 15 (45.5%) | 3 (6.8%) |

**FN = false-negatives SF = secondary findings**
**Results are presented as number and percentage (%) of total in parenthesis. Distinction was made** **between non-annotated conclusions although containing SF and those with a deficit between the** **number of SF present and annotated. The percentage of total conclusions with SF was calculated.**

**Table 7.**
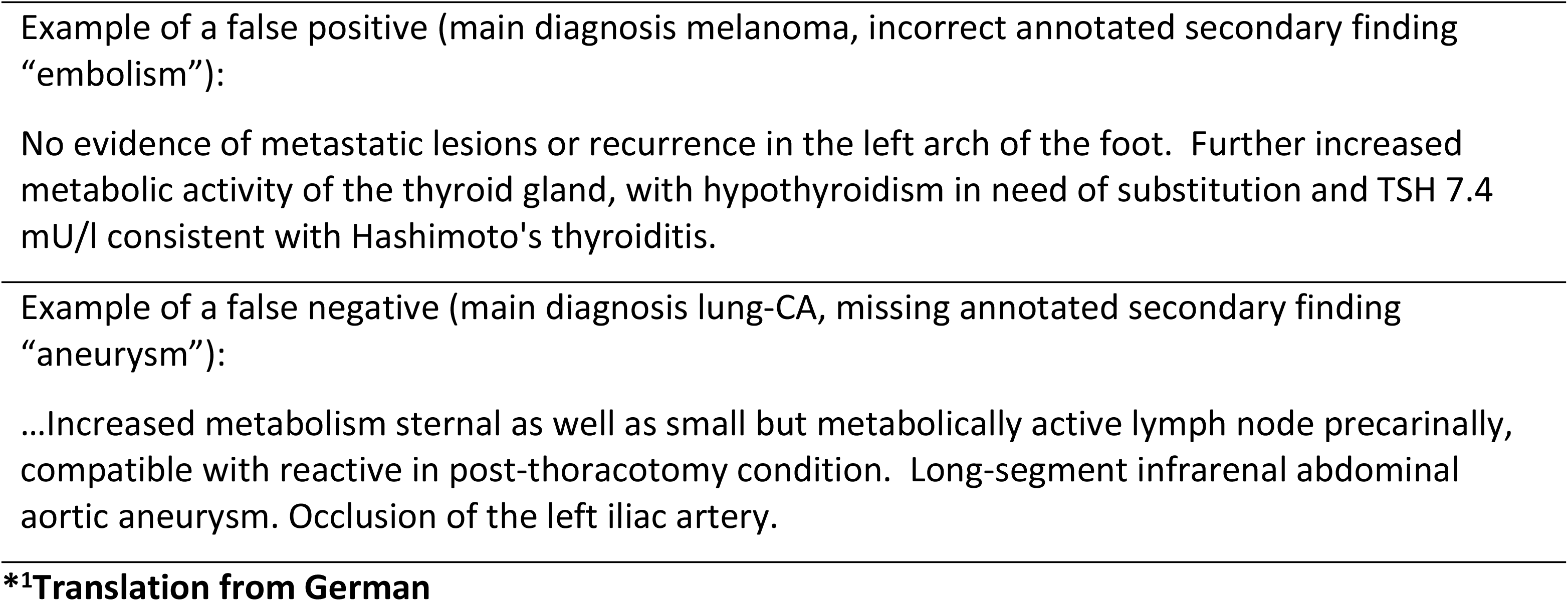
Example of a typical case of a false positive and false negative radiological conclusion *^1^, respectively.

**Figure 2.**
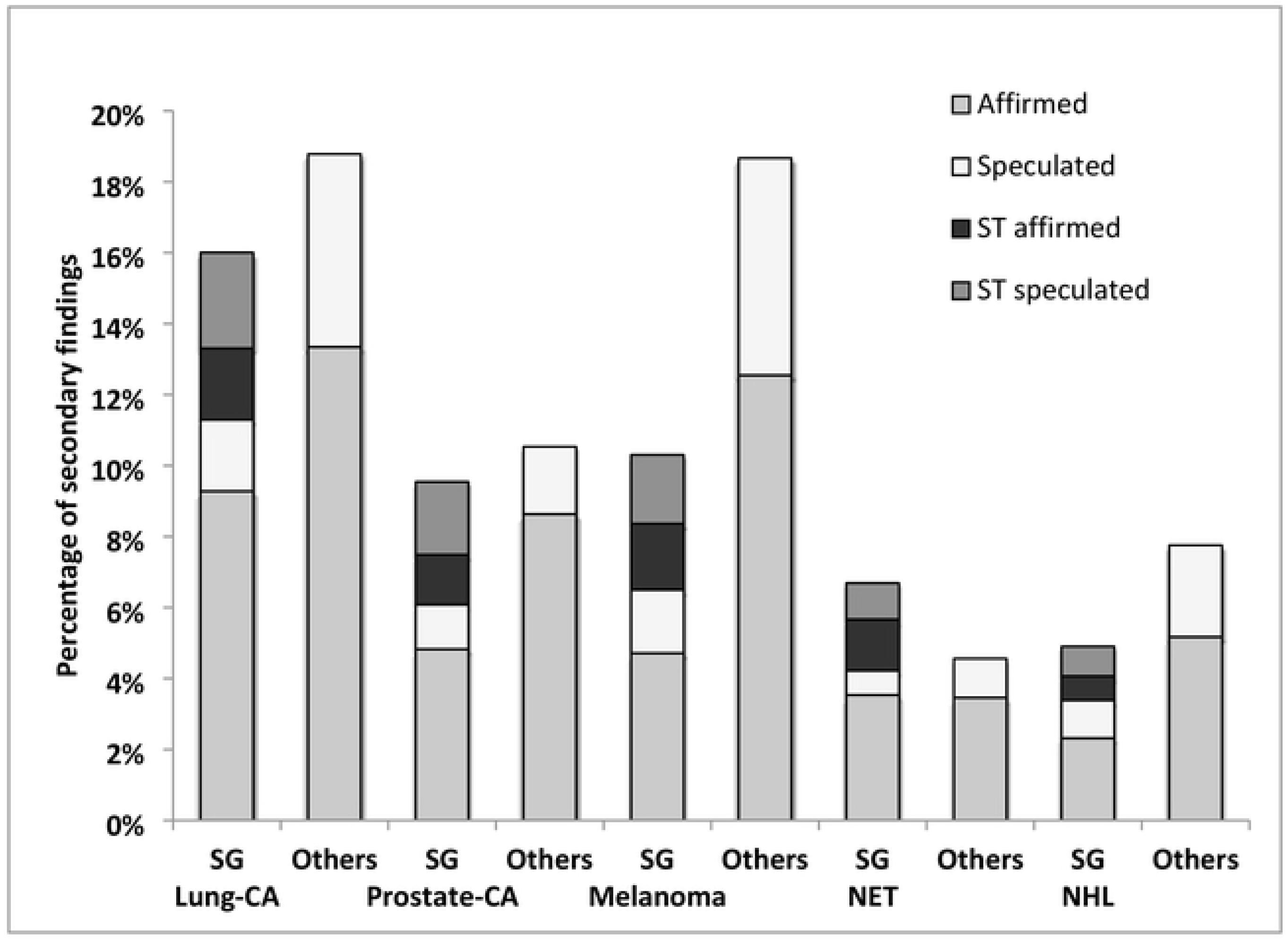
Chart illustrating the pattern of affirmed and speculated subgroups (SG) of secondary findings and “others” (conglomerate of unspecific subgroups) as identified by the NLP system. Secondary tumors (ST) are a special part of subgroups.

#### False positives and false negatives in secondary findings

In order to classify the accuracy of the NLP-System, the number of false positives and false negatives were also evaluated.

In cases with the main diagnosis NET the highest error rate of false positives was found. 17 out of 295 secondary findings were rated as false positives which results in an error rate of 5.8%. In contrast, hardly any false positives were found in the cohort diagnosed with NHL. There only 1% of all secondary findings were false positives and there was no incorrect assignment to affirmed or speculated. Overall, the rate of false positives (1.0% - 5.8%) and incorrect assignment (0% - 1.1%) was very low compared to the overall rate of annotated diagnoses. The complete calculation is listed in Table.

Error rates of false-negative secondary findings calculated using a random sample of 100 cases per principal diagnosis ranged from 6.1% (NET) - 24% (prostate-CA) meaning that there were up to 24% of conclusions with a secondary diagnosis that was not found. More frequently, in conclusions with multiple secondary findings not all of them were recorded. This error rate varied from 6.8% for NHL to 45.5% for NET. In part, this high number can be attributed to the fact that multiple secondary diagnoses were sometimes not found in a single conclusion. The complete calculation is listed in Table.

One example each of a false positive and false negative case is shown in Table.

## Discussion

This study evaluated the applicability of an NLP technique for the annotation of secondary findings from free-text written German radiology conclusions. Furthermore, the annotation results were interpreted to discuss them in the context of five main oncological diagnoses.

The gold standard for the evaluation from radiology reports currently still is the manual selection of information by experts. However, this is very time-consuming. Our data show that NLP technology is a useful tool to efficiently extract secondary diagnoses from German freehand written radiology texts in a time-saving manner. Since the clinical significance of secondary diagnoses varies considerably between different patient groups, being able to extract them quickly and reliably from radiology reports is important for quality management.

In identifying the main diagnosis within the clinical information, we achieved excellent F1-scores between 0.65 and 0.95 without specific training. demonstrating the efficacy of the NLP algorithm. The positive predictive value was between 0.96 and 1, indicating that all diagnoses found were correct. Merely the sensitivity could be improved in cohorts with NET and lung-CA by training the NLP-System (24), since currently up to 50% of the diagnoses are still hidden for the algorithm. However, the complexity of the German language also plays a significant role here, as there are a large number of paraphrases and synonyms in our freehand written clinical information for these two types of cancer. It has already been shown in other studies that complex non-English texts from the German language family can achieve very good scores in all three metrics by training the NLP-algorithm (32, 33). However, achieving perfect quality is often challenging and may not be necessary for large data sets.

Annotation of the secondary diagnoses was done in three steps. First, all annotated secondary diagnoses were grouped into supersets, then these were subdivided into subgroups and “others”. As a final step, the false positives and false negatives were identified.

Among the supersets the most affirmed and speculated secondary findings were found in patients with a principal diagnosis of lung-CA. The frequency and classification of the clinical relevance of secondary findings in lung-CA is very heterogeneous in the literature and ranges from 7 - 27% (34). In the evaluation of this current study, the high rate of mechanical disorder was particularly striking. This includes, for example, secondary findings such as atelectasis and pleural effusion, which are typically more common in lung-CA and its treatment (35) than in other oncologic diseases. In all other cohorts, the most common secondary findings were also found in the superset of mechanical disorder.

Secondary tumors are a special part of superset which was not further divided into smaller subgroups. These have been affirmed second most frequently in all cohorts. Again, the number was highest in the cohort with lung-CA. In a previous study, a secondary tumor was found in 12.6% of patients with a primary diagnosis of lung-CA (36). Secondary malignancies are rather rare incidental findings (37), but can have a significant impact on therapy if confirmed. In our study, mainly benign secondary tumors like adenomas of the adrenal gland were identified as secondary tumors.

The largest number of “infectious or inflammatory disorders” was found in the cohort of melanoma patients. A more detailed classification of this subset into subgroups shows that these are mainly cases of sinusitis.

The lowest number of secondary findings was found in the cohort with NHL and NET. In part, the number of secondary findings may be explained by the type of therapy. Since many neuroendocrine tumors are treated primarily with surgery and specific drugs (38), the full-body impact and thus secondary findings are comparatively less than in patients with melanoma or lung-CA.

Melanoma patients in contrast often receive immunotherapy, which increases the risk of infectious diseases and patients with NHL are receiving immunochemotherapy, which weakens the immune system (39). Patients with prostate-CA are the oldest cohort with an average age of 70 years. At this age, people frequently have other concomitant diseases by nature and therefore some secondary findings were also found in further studies (40). Earlier studies have shown that some secondary findings can have a significant impact on therapy (41, 42). Therefore, it is very important to be able to extract this information reliably and quickly in order to adapt patient management if necessary.

The rate of false-positives was very low. Some false positives could be prevented by training the system slightly more (24). Sometimes related terms were recognized as diseases by the NLP system (e.g. lymph node metastases as lymphoma) or confused (e.g. ectasia of the aorta as hydronephrosis). Abbreviations and their ambiguity can also be a problem. For example, by partially interpreting the abbreviation “ALL” (acute lymphoblastic leukemia) as “all” some false positives were generated.

The matching of the secondary findings to the concepts affirmed and speculated succeeded almost without error. Any confusion occurred only due to linguistic inaccuracies or hints hidden in sentences within the conclusions. Concepts in radiological reports that are interpreted differently even by clinicians have already been identified in a previous study (43).

The rate of false negatives was actually higher than the number of false positives. This can also be attributed to the lack of training on the one side. Some false negatives are due to language diversity in the conclusions. Besides many synonyms, there are also many expressions in the German language that have the same meaning. Some errors are due to ambiguity or false negation detection (44). In a previous study (45), the number of false negatives was also higher than the number of false positives. Here, language recognition errors, syntax errors, or the inability to recognize the plural of a word, among others, were identified as sources of error. Many false-negative errors could be resolved by standardizing radiology reports (46). Another study (9) also recognized that shorter reports lead to fewer errors in NLP recognition. The higher amount of information in more detailed reports could negatively affect NLP detection.

In summary, NLP is a useful tool for extracting clinically relevant data, such as secondary findings, from radiology reports. This is important because no statistics are available yet regarding the most common secondary diagnoses in patients with particular oncologic diseases. Furthermore, an NLP tool can help to prevent clinicians from missing important information and to save time in the evaluation process. This can also be used to extract important information from medical reports that otherwise would require tedious re-reading. Since most NLP systems are specialized for English texts or certain text types, they have to be trained for other applications (47). However, free-text written radiology reports are in some ways also a challenge for NLP, since natural language also uses ambiguous terms that are difficult to classify by an automated system, but which an expert may easily infer by understanding the context. Therefore, free texts are supported by further machine learning processes in some studies (48). On the other hand, even experienced investigators might misinterpreted free-language reports authored by colleagues (49). Thus, there is a need for standardization. NLP technology could be helpful to develop improved imaging reporting in radiology and nuclear medicine.

## Conclusion

NLP technology can be used to efficiently and easily extract important data retrospectively from radiology texts. Thus, NLP is a helpful tool for research and patient management. The complexity of human language and the resulting difficulties for NLP technology should be considered when writing the respective reports.

## Data Availability

All relevant data are within the manuscript and its Supporting Information files. The datasets generated and analyzed during the current study are not publicly available due to sensitive information of patients but are available in anonymous form from the corresponding author on reasonable request.

## Abbreviations

PET/CT: Positron emission tomography/computed tomography
NLP: Natural language processing
AI: Artificial intelligence
RadLex: Radiological Lexicon
ICD: International Classification of Diseases
NHL: Non-hodgkin-lymphoma
lung-CA: Lung cancer
prostate-CA: Prostate cancer
NET: Neuroendocrine tumor
SF: Secondary findings
SG: Subgroups
ST: Secondary tumors
FP: False-positive
FN: False-negative

## Declarations

### Ethics approval and consent to participate

This study was approved by the Ethics committee of the University of Tuebingen, reference number 064/2013B01. Written informed consents were waived due to retrospective nature.

### Consent for publication

Not applicaple.

### Availability of data and material

The datasets generated and analyzed during the current study are not publicly available due to sensitive information but are available in anonymous form from the corresponding author on reasonable request.

### Competing interests

BK is an employee of Empolis Information Management GmbH (Kaiserslautern, Germany). The other authors declare no conflict of interest.

### Funding

No funding has been received for this publication.

### Authors′ contributions

SG originated the idea for the project. JS extracted the conclusions needed. AD took care of data privacy issues in exporting data. BK performed the annotation and assisted in interpretation. JS and SG analyzed the data. CR summarized methods for generating radiological reports. JS prepared the figures and wrote the first draft of the manuscript. BG, CP, and HD co-wrote the final version of the manuscript and were significantly involved in advising on clinical aspects. All authors read and approved the final manuscript.

## Acknowledgements

Not applicable

## Notes

### Competing Interest Statement

I have read the journal's policy and the authors of this manuscript have the following competing interests:BK is an employee of Empolis Information Management GmbH (Kaiserslautern, Germany). The other authors declare no conflict of interest.

### Funding Statement

The author(s) received no specific funding for this work.

